# The Effects of Improved Housing on Malaria Transmission in Different Endemic Zones: A Systematic Review and Meta-Analysis

**DOI:** 10.1101/2023.08.06.23293581

**Authors:** Mukumbuta Nawa, Olatunji Adetokunboh

## Abstract

**Introduction:** Improved housing has been shown to reduce the risk of malaria infections compared to traditional houses; however, it is unclear if the effects differ in different malaria transmission settings. This study evaluated the effects of improved housing on malaria transmission among different endemic areas.

**Methods and Analysis:** Electronic databases, clinical trial registries and grey literature were searched for randomised controlled trials, cohort studies, case-control studies, and cross-sectional surveys on housing done between 1987 and 2022. Forest plots were done, and the quality of evidence was assessed using the Grading of Recommendations, Assessments, Development and Evaluation Framework.

**Findings:** Twenty-two studies were included; twelve were cross-sectional, four were case-control, four were cohort studies and two were RCTs. RCTs indicated that modern houses did not protect against malaria compared to traditional houses. Cohort studies showed an adjusted risk ratio of 0.68 (95% CI 0.48 – 0.96) and Cross-sectional studies indicated an adjusted odds ratio (aOR) of 0.47 (95% CI 0.31 – 0.72). By endemic transmission regions, the adjusted odds ratio in the high endemic was 0.43 (95% CI 0.29 – 0.63); in the moderate transmission regions, aOR = 0.91 (95% CI 0.91 – 1.07) and in the low transmission settings, aOR = 0.42 (95% CI 0.26 – 0.66).

**Conclusion:** The evidence from observational studies suggests that the risk reduction associated with modern housing is comparable or higher in low malaria transmission settings compared to high transmission settings. Evidence from RCTs in high-transmission settings shows that house improvements may induce risky behaviours such as staying outside for longer hours.

**Key Messages:**

1. It is known that improved housing reduces the risk of malaria compared to traditional housing; however, the effects of improved housing in different endemic settings are unclear.
2. To the best of our knowledge, this is the first time a systematic review and meta-analysis has stratified the effect measures of improved housing on malaria transmission in different transmission settings.
3. Our study found no literature from high-quality research designs such as RCTs and Cohort studies on improved housing in low and moderate transmission settings. We call on researchers and funders to conduct and support high-quality research designs in low and moderate-transmission areas, especially in Africa, as more countries are reducing their malaria burdens due to increased interventions. This will help to achieve and sustain malaria elimination.
4. Piecemeal improvements, such as closing eaves, screening and iron roofing, are not necessarily associated with a reduced risk of malaria. They may induce risky behaviours due to poor ventilation and higher indoor temperatures resulting in residents staying longer outside thus exposing themselves to infective mosquito bites.

## INTRODUCTION

The fight against malaria has stalled in recent years partly due to the emergence of resistance to insecticides used in Long-Lasting Insecticide-treated Nets (LLINs) and Indoor Residual Spraying (IRS), reduced investments and disruptions in interventions during the Coronavirus Disease of 2019 (COVID-19) pandemic [1, 2]. Researchers have called for novel tools to enhance the fight against malaria for global eradication to be realised in line with the WHO Global Technical Strategy of eliminating 90% of incident cases and deaths by 2030 [3]. In 2021, the WHO approved a malaria vaccine called RTS, S/AS01; however, this presents some logistical challenges, such as huge investments required in the roll-out [4]. Others have called for community and human-centred approaches [5].

While the disease is raging on, researchers and policymakers are looking for solutions to emerging challenges. The case for housing infrastructure improvements in the fight against malaria, which was superseded by the discovery of chemical agents, has emerged [6–8]. In sub-Saharan Africa, traditional housing structures are typically stick-and-mud walls and grass-thatched roofs [9]. These traditional houses have open eaves and holes in the walls to allow for ventilation; however, flying insects such as mosquitoes can also freely enter and exit them.

On the other hand, modern housing structures have brick and iron/tiled roofs. Studies have shown that the risks and odds of malaria infection can be reduced by about 47% for those who dwell in modern houses compared to those who dwell in traditional houses [9–11].

While there is sympatry or co-existence of primary vectors, the primary *Anopheles* mosquito vectors that predominantly transmit malaria in highly endemic areas differ from those that predominantly transmit malaria in low endemic areas in terms of their feeding host preferences, resting behaviour, Entomological Inoculation Rates (EIR) and Sporozoite Infection Rates (SIR) [12–17]. The effect of barrier prevention methods such as housing structures is likely to differ for high-endemic areas compared to low-endemic areas. Our study, therefore, addressed this knowledge gap and can help government agencies target effective policies and interventions relevant to local settings.

## METHODS AND ANALYSIS

We used the Preferred Reporting Items for Systematic Reviews and Meta-Analysis (PRISMA) guidelines to prepare a systematic review and meta-analysis [18]. The protocol was registered with the Prospective Register of Systematic Reviews (PROSPERO – ID 357186).

### Study Settings

This review included studies from sub-Saharan Africa, South America, and Middle and East Asia and was stratified according to malaria-endemic zones.

### Inclusion and Exclusion Criteria

#### Types of Studies

Studies included were RCT designs and observational studies such as cross-sectional surveys, case-control and cohort studies published between 1987 and June 2022 in line with the establishment of the Roll Back Malaria Initiative in 1987. All studies with clear effect measures were included, whilst those that were qualitative or without effect measures were excluded. Those without clear geographical areas where the studies were conducted were also excluded.

#### Type of Participants

We included studies that compared malaria occurrence in all types of residents, whether children under five years or adults or specific subsections of adults such as pregnant women.

#### Interventions

Studies had to be clear that they compared modern housing structures against traditional or non-standard housing structures. Modern housing structures had brick walls, iron/tiled roofs, ceilings, and closed eaves, while traditional houses were made of mud or grass walls and grass-thatched roofs with open eaves.

#### Type of Outcome Measures

Different studies measure malaria outcomes in different ways. Cross-sectional studies measure malaria prevalence diagnosed by blood slides using light microscopy; in this case, participants can either have clinical symptoms or be asymptomatic. Cohort studies and RCTs measure malaria incidence. Our study included prevalence and incidence as outcomes and analysed them separately by different endemic areas.

#### Information Sources

We searched major databases for peer-reviewed journal articles on the subject, including Cochrane, MEDLINE (PubMed), Scopus, The Global Index Medicus and Web of Science. Peer-reviewed scientific conference proceedings, such as the American Society of Tropical Medicine and Hygiene, and The International Congress for Tropical Medicine and Malaria, were searched. Further, we also searched clinical trial registries, including the WHO clinical trials registry and the American clinicaltrials.gov and grey literature.

#### Search Strategy

A literature search strategy was developed in Medline using Mesh subject headings combined with free text. The search strategy developed in Medline was adapted to other databases in collaboration with the University of Stellenbosch librarian and has been attached as supplementary material.

#### Study Records

The identified articles were imported into a citation reference manager called Endnote; Endnote was used to de-duplicate articles.

#### Screening for Eligibility

Rayyan QCRI Software was used to screen the articles for eligibility [19]. Three reviewers (NM, CMM and SBT) independently screened the titles and abstracts in Rayyan. Disagreements were resolved by discussion between the team members. (CMM and SBT) then read the full text for the selected articles and finalised the screening process with NM. OA supervised the screening process.

#### Data Extraction

Two reviewers (CMM and SBT) extracted data from selected studies into a pre-piloted data extraction form. The consensus was established between the two, and arbitration by the third reviewer (NM) when needed. The data points included: authors, year of publication, sample size, study design, effect measures with 95% confidence intervals, type of participants, and geographical coverage.

#### Assessment of Risk of Bias in Included Studies

Three reviewers (NM, CMM and SBT) assessed the risk of bias in the studies in duplicate. The first was for RCTs using the Cochrane Risk of Bias tool (RoB-2), which assessed selection, performance, detection, attrition, reporting, and any other bias [20]. The ROB-2 Tool categorised the quality of results based on the algorithm into low risk, some concerns, or a high risk of bias. The risk of bias for observational studies was assessed using the Risk of Bias for Non-Randomised Studies for Exposure (RoBINS-E) [21]. The risk of bias in the papers was reported as low risk, moderate risk, serious or critical risk based on the algorithm.

#### Measures of Treatment Effects and Associations

The outcome of this study was to establish the effects and measures of the association of modern houses on malaria cases (incidence and prevalence) stratified by low, moderate, and high endemic settings. Clinical trials and cohort studies that report risk ratios were analysed and reported separately. At the same time, prevalence and case-control studies that report odds ratios were also analysed and reported separately.

#### Unit of Analysis Issues

For follow-up studies such as RCTs and Cohort studies, we used the Incidence Risk Ratios (IRR), Rate Ratios or Absolute Risk Differences to compare malaria incidence in standard compared to traditional houses in different endemic settings and age groups. Where events occur below 10% in the samples, odds ratios were used as they are better estimates in rare events. In cross-sectional and case-control studies, the analysis unit used was odds ratios.

#### Assessment of Heterogeneity

In line with the Cochrane guidelines, heterogeneity in the studies was assessed using the *I^2^* statistics in the meta-analysis, which is calculated by:

I^2^= ((Q-df)/Q) *100, where Q is the Chi^2^ and df is the degree of freedom.

An *I^2^* of 75-100% would be interpreted as considerable heterogeneity, 50-90% as substantial heterogeneity, 30-60% as moderate, and below 40% as unimportant [22].

#### Assessment of Reporting Biases

We planned to assess reporting bias using funnel plots where we had at least ten studies included in the meta-analysis and to look at the symmetry of the funnel plots.

#### Data Synthesis

A summary of how many articles were identified during the literature search, how many were excluded at what stage of the process, why they were excluded, and how many were finally included are presented in a flow diagram [23]. A descriptive table of included articles, where, when, authors, and effect sizes are presented. We made forest plots of the analysis displaying pooled effect measures, 95% confidence intervals, p values, Chi-square, and *I^2^* values. We conducted meta-analyses of similar studies to find the pooled effect measures by endemic zone using RevMan for Windows (version 5.4) [24]

Similar study designs that reported the same measures of association and effect measures were used to create separate forest plots. We run separate forest plots for each study design using the reported effect measure, whether risk ratio, rate ratio, absolute risk difference, or odds ratio low, moderate, and high endemic settings.

#### Certainty of the Evidence

The certainty of the evidence was assessed using the Grading of Recommendations, Assessment, Development and, Evaluations (GRADE) framework [25]. Evidence was categorised as very low, low, moderate, and high quality. We included the high-certainty findings in the GRADE table.

#### Ethical Considerations

The Department of Global Health research protocol panel at the University of Stellenbosch reviewed and approved this study. We also obtained an exemption for review from the Health Research Ethical Committee at the University of Stellenbosch as it does not involve human subjects (HREC Reference number: X22/08/020).

## RESULTS

A total of 3167 articles were collected from the database search, and an additional three from grey literature, totalling 3170. Following screening, 2923 articles were excluded, and a full-text screening was done on 247 articles. A total of 141 were excluded on full-text screening, and 84 were excluded because they compared components and not comprehensive houses. Figure 1 shows the inclusion flow chart.

### Included Studies

The majority of the studies included were cross-sectional study designs 12 (54%), case-control studies 4 (18%), cohort studies 4 (18%), and RCTs 2 (9%). More than three-quarters of the studies were done in Africa 19 (86%), less than a fifth in Asia 3 (14%) and none in Latin America. Over half of the studies were cross-sectional surveys, and less than 10% were Randomised Controlled Trials implying low levels of high-quality research in malaria. These differences were statistically significant (P value < 0.001).

### Study Settings

The majority of the studies done in Africa were done in high endemic settings 10/19 (53%), a third 6/19 (32%) from moderately endemic settings and 16% (3/19) in low endemic settings. Those from Asia were from moderate endemic settings in India and Pakistan.

### Characteristics of Study Participants

Among the studies included those that assessed malaria parasites among participants were 20 studies, and altogether, there were 141 242 participants. The participants included children aged 0 – 15 years who were 44 785 (31.7%) in 9 studies; the larger proportion was the general population who constituted 95, 785 (67.8%) in 11 studies and only one study compared malaria incidence among pregnant women in traditional and modern houses involving 753 pregnant women. Further, two entomological studies were included in which 129,508 mosquitoes were captured to assess the human biting rates and vector densities in traditional houses compared to modern houses. Table 1 gives a summary of the characteristics of included studies.

### Characteristics of Interventions/Exposures and Comparisons

All 22 studies included in this review compared modern houses against traditional houses; these traditional houses ranged from mud or grass wall and grass thatched roofed houses in Africa to stick hamlets and bamboo or wood walls and thatched roofs in Asia. Modern houses being compared were made of brick walls and iron/tiled ceramic roofs.

### Primary Outcomes

The primary outcomes reported in the included studies were malaria prevalence and incidence depending on the study design. A total of 17 outcomes of the interventions reported the prevalence of malaria parasites in the respondents. A total of 4 outcomes were reported on malaria incidence. Some studies reported more than one outcome.

### Secondary Outcomes

Three entomological studies reported indoor resting vector densities, while the other three reported human biting rates. No entomological study among the included studies linked entomological inoculation rate and housing structures.

### Excluded Studies

A total of 84 studies were excluded on the basis that though they had effect measures on housing structures comparing malaria in traditional versus modern structures, however, they only compared components of houses such as thatched roof versus iron/tiled roof, ceiling versus no ceiling, closed eaves versus open eaves or mud walls versus brick walls.

### Risk of Bias Assessment

Out of the 22 studies included, two were randomised community trials [26, 27] and were assessed for risk of bias using the Cochrane Risk of Bias Tool [20]. The two open-label community trials had one weakness i.e. not blinding either the participants or the field assessors; however, the evaluators who did the lab tests and analysed the data were blinded, thus minimising the risk of bias. Further, the allocation sequence was computer generated from the census sampling frames; thus, the risk of bias for both studies was judged as some concerns by two assessors who worked independently. The summary of the risk of bias in RCTs is in Table 2.

Twenty of the included studies were observational and were assessed for the risk of bias using the Risk of Bias in Non-Randomised Studies of Exposure (RoBINS-E) Tool [21]. Five of the studies had a serious risk of bias arising from recall bias due to prolonged periods assessed [28, 29], risk of selection bias [30] and confounding due to the use of unadjusted odds ratios in the studies [31, 32]. Ten included studies had moderate concerns, mainly arising from residual confounding in cross-sectional and case-control studies, even after multivariate regression adjustment. Four had a low risk of bias mainly because they were cohort studies [33–36]. A Summary of the risk of bias assessment for the observational studies is shown in Table 2.

### Effects and Associations of Interventions/ Exposures on Outcomes

The overall association of modern houses on the risk of malaria parasitaemia compared to traditional housing among cross-sectional surveys using the adjusted odds ratios reported in the individual studies was a reduction in the adjusted odds ratio of 0.47 (95% CI 0.31 – 0.72). The overall heterogeneity was high at I^2^ = 79% and was statistically significant (P value < 0.001). The within-groups heterogeneity was low and not statistically significant; however, heterogeneity was high at 89.6% between groups, and this was statistically significant (P value <0.001), implying significant differences in the association of modern housing in different malaria transmission settings. Table 3 summarises the pooled measures of associations.

When the effect of modern housing was stratified by endemicity, the effect in the high endemic zones was at odds ratio 0.43 (95%CI 0.29 - 0.63) and was statistically significant. The heterogeneity in the high endemic zone was low at I^2^ = 0.39% and not statistically significant, indicating agreement in the general direction and magnitude of the odds ratios among the cross-sectional studies.

The association of modern housing in the moderate malaria endemic zones compared to traditional housing was found not significant at odds ratio 0.91 (95%CI 0.77 - 1.08). There was only one study done in India [28]. In the low endemic zone, there were only two studies, one from Zambia and the other from eSwatini(Swaziland); the association of modern housing compared to traditional housing showed an odds ratio of 042 (95%CI 0.26 – 0.66, I^2^ = 1% P value = 0.32) which was comparable to the high transmission settings probably because all studies were done in Africa.

We further conducted a meta-analysis of cross-sectional studies using unadjusted odds ratios from reported actual numbers of infections and total participants included in studies. The pooled measure of association was an odds ratio of 0.77 (95%CI 0.55 – 0.81), and the I^2^ was 0%. This association was less than the one calculated from adjusted odds ratios, probably because of confounding from other factors that were not adjusted for and, therefore, may not be reliable.

The study further assessed the associations of modern housing compared to traditional housing using case-control studies that reported adjusted odds ratios by endemic zones. There were only two studies in the meta-analysis, one done in Zambia and the other in northern Namibia. The overall effect of modern housing compared to traditional housing was an odds ratio of 0.52 (95%CI 0.38 – 0.70, I^2^ = 0%, P value = 0.40) which shows that modern housing had a statistically significant effect in reducing the risk of malaria compared to traditional housing. When stratified by endemicity, only one case-control study was included in the high endemic region, and the effect measure was an adjusted odds ratio of 0.33 (95%CI 0.11 – 0.99). There were no studies included which were done in the moderately endemic regions.

In contrast, one study was included in the low malaria endemic region, and the effect measure was an odds ratio of 0.54 (95%CI 0.39 – 0.74). Due to the few studies included in the meta-analysis, the heterogeneity was low.

Our review further analysed case-control studies that reported an unadjusted number of malaria infection events against totals and conducted a meta-analysis. Only two studies were included, one from Egypt and another from Zimbabwe, which were both in low transmission settings. The pooled measure of association was an odds ratio of 0.33 (95%CI 0.06 – 1.75, I^2^ = 71% and P value = 0.06). This association was not statistically significant because of a wide confidence interval, few studies, and likely confounding from the unadjusted odds ratios used.

Further analysis of observational studies was done using cohort studies that compared adjusted incidence (risk) ratios among residents of modern houses against traditional houses. Two cohort studies compared the adjusted risk ratios in modern houses to traditional houses, both done in Uganda [33, 35]. There was a reduced Incidence Risk Ratio (IRR) of 0.68 (95%CI 0.48 – 0.96, I = 71%, P value = 0.06). The risk reduction was statistically significant based on the confidence intervals. Still, the heterogeneity was not significant, probably because of the few studies and that they were done in the same country.

A meta-analysis of cohort studies that reported unadjusted Incidence Risk Ratios found only two studies from Uganda and pooled risk ratios of 0.89 (95%CI 0.70 – 1.14). This effect was not statistically significant, unlike the ones done from the same country Uganda in similar settings that reported adjusted Risk Ratios.

The same two cohort studies done in Uganda that reported non-significant risk ratios also reported unadjusted odds ratios, which were statistically significant (Risk Ratio 0.63 (95%CI 0.41 – 0.97) when pooled in a meta-analysis.

In cohort studies, we further explored the association of modern housing compared to traditional housing using mosquito vectors’ Human Biting Rate (HBR). Two studies from Uganda reported the unadjusted risk ratio using HBR (RR 0.53 (95%CI 0.43 – 0.65) [34, 37].

We further included two randomised community trials (RCTs) in this review; however, they could not go into the meta-analysis because of the different effect measures used, one reported on vector density mean ratio [26], and the other reported on IRR of malaria incidence and vector densities [27]. If the house is an ordinary modern house with closed eaves without an additional screening of gables and special fitting of doors, the mean ratio of vectors indoors is increased to 2.99 (1.96 – 4.57) compared to traditional housing; however, if modern houses are screened with gables and well ventilated, and doors are specially fitted, the mean ratio of *Anopheles* gambiae was reduced to 0.06 (0.03 – 0.10) [26]. Another RCT found that modern/improved housing did not protect against clinical malaria; the IRR for clinical malaria in modern houses was 1.68 (95%CI 1.11 – 2.55), while the IRR for vector density for *Anopheles* gambiae in the modern house was 1.28 (95%CI 0.87 – 1.89) compared to traditional houses.

### Quality of Evidence (GRADE)

The studies included in this review provide moderate evidence from RCTs and cohort studies and low to very low evidence from cross-sectional and case-control studies. [27]. Table 4 shows the summary table for the certainty of evidence using the GRADE Approach.

## Discussion

This systematic review and meta-analysis sought to find the effects and measures of association between housing and malaria infections in different malaria endemic zones. Previous meta-analyses, particularly non-Cochrane studies that included sufficient observational studies, found high heterogeneity in the measures of associations between housing structures and malaria parasitaemia [38]. The study found mixed results, particularly in RCTs. One study did not find additional protection from improved housing; in fact, that study found that improved houses increased the risk of malaria incidence and indoor vector densities compared to traditional houses [27]. The reasons for increased risk were related to increases in temperature in modern houses with Iron roofs, so people opened doors allowing for ventilation, but mosquitoes also entered. The people also stayed longer outdoors, thus exposing themselves to mosquito bites. The other study also found an increased risk of malaria and indoor densities in improved houses if they were not specially fitted with screens and closures of not only eaves but gobbles and doors specially fitted [26]. These RCTs were both done in one country, the Gambia, which has a high coverage of other interventions such as IRS and LLINs. The region also has low to moderate malaria transmission, so the effect of modern housing in high transmission settings from RCTs was not covered due to a lack of studies. Other existing RCTs on improved housing covered components of improved housing such as screening and closing of eaves, iron roofs and brick walls [6, 39–42]. In these and other similar studies, various piecemeal improvements to housing components were tested for acceptability and associations or effects of these improvements, demonstrating some reduction in malaria risks [6, 9, 10, 39]. As seen from the Gambian RCTs, intervening with components only such as closing of eaves, iron roofing may have other unintended consequences such as poor ventilation, increase in indoor temperature and carbon dioxide which may also modify human behaviour leading to more risk of exposure to malaria [26, 27]. We therefore advocate for holistic interventions that will not only address infrastructure components, but also human behaviour associated with the modifications or improvements.

The majority of observational studies, on the other hand, such as cohort, case-control and cross-sectional studies in our systematic review and meta-analysis, seem to suggest that comprehensive house improvements that include both iron roofs and brick walls reduce malaria risk and indoor vector densities with very few showing that the measures of association are not statistically significant [31, 34, 37, 43, 44]. However, socio-economic factors such as wealth, education, nutritional status and health status are also associated with living in improved houses compared to living in traditional houses [45, 46]. Therefore, even when some socio-economic factors were adjusted for in the observational studies included in our review and meta-analysis, residual confounding was still an important factor and as such, the results were considered low to very low certainty of evidence using the GRADE system. They must be interpreted with caution [25].

In terms of the effects of housing in different endemic settings, using the high to moderate-quality evidence from RCTs, we did not find sufficient studies in all transmission settings to compare the effects in different endemic settings using like-to-like. The two RCTs we included were done in the Gambia, categorised as moderate endemic settings [26, 27]. Four cohort studies were all done in Uganda and were categorised as high-endemic settings [33–36]. Of the four case-control studies included, three were in low transmission settings [31], and only one was in high endemic settings [11]. So the comparisons could only be done using cross-sectional studies where there were studies in all endemic settings and allowed us to do comparisons using the same measures of association. Using unadjusted odds ratios which are very low-quality evidence studies due to the risk of confounding, the risk reduction of modern housing seemed to have been higher in low and moderate settings compared to high endemic settings. However, using cross sectional studies which reported adjusted odds ratios in all settings, the measures of association were comparable in low and high transmission settings. One study from India showed a very minimal risk reduction which was not statistically significant, probably because it only measured malaria in the adult population aged 45 years and above, which is different from children aged below five years and the general population, which most cross-sectional studies in Africa measure malaria in [28].

From an entomological perspective, high transmission settings have higher entomological inoculation rates (EIRs), so people get bitten many times by infected mosquitoes, and you would expect residents of traditional houses that do not impede mosquito entry to have a higher probability of infections compared to people in modern houses [47]. Conversely, as the EIRs reduce in moderate and low endemic transmission settings, you would expect a dose-response-like effect of reduced measures of association in moderate and low endemic settings. However, emerging evidence from this study using adjusted odds ratios suggests similar associations between low and high transmission settings, whilst unadjusted odds ratios show more risk reduction in low-endemic settings. So based on the dose-response-like associations using higher EIRs in higher endemic settings and lower EIRs in moderate and low malaria endemic settings, we would have expected the measures of association of modern housing and risk of malaria to have been higher in high endemic settings compared to low endemic settings. Something else, therefore, needs to explain why our findings seem to suggest comparable to higher measures of association in the low endemic settings compared to higher endemic settings.

From an immunological perspective, those that get bitten more times in the higher transmission settings develop acquired immunity and can fend off infections and clinical disease even when bitten by infected mosquitoes multiple times [48]. Conversely, those in low transmission settings may not have had frequent bites enough to confer acquired immunity; for example, the EIR in some places in Uganda may be as high as 310 infective bites per person per year, whilst in low transmission settings such as Botswana, Namibia and the Southern parts of Zambia, the EIRs are below 1.6 infective bites per person per year [49, 50]. A person bitten by an infective mosquito less than twice a year is less likely to develop acquired immunity than another who gets bitten by infective mosquitoes 310 times a year. This can explain why the measures of association seem to suggest a comparable to higher risk reduction in low transmission settings compared to higher transmission settings because the population in low transmission settings may have low acquired immunity to malaria. So, a barrier method like improved housing may prevent the few bites from happening in improved houses compared to traditional houses that do not offer mosquito bites protection. We call upon more research using better designs, such as RCTs and cohort studies in different endemic settings, to elicit higher-quality evidence to better inform policymakers and program managers.

Elsewhere, policymakers and program managers of malaria programs have noted the reduced effects and associations of other interventions, such LLINs and IRS, in low transmission settings[51, 52]. Despite the low to very low quality of evidence available, the findings of this study may therefore be of interest if, indeed, modern houses have higher or comparable effects and associations in low transmission settings compared to high endemic settings. Improved housing to modern standards can be an addition to the tools available in the fight against malaria in low and moderate settings, especially now as we garner towards malaria elimination by 2030.

### Conclusion

The currently available evidence on measures of association and effects of complete house improvement on malaria transmission in different endemic transmission settings is limited to low and very low-quality evidence. The evidence suggests that the risk reduction associated with modern housing compared to traditional housing structures is comparable or higher in low malaria transmission settings compared to high transmission settings. Evidence from high and moderate-quality studies done in high-transmission settings shows that complete house improvements may have benefits of reducing the risk of malaria transmission and indoor vector densities. However, the improvements in housing structures may induce risky behaviours such as staying outside for longer hours if the improvements to housing lead to increased indoor temperatures and poor ventilation. Piecemeal improvements to housing, such as closing eaves, screening and iron roofing, do not necessarily result in a reduced risk of malaria; rather, holistic interventions that consider conditions such as house ventilation, temperature and human habits may be more beneficial.

### Limitations

The main limitation of this study was that there were few high and moderate-quality evidence studies, such as RCTs and cohort studies in all endemic settings, so it mainly relied on low to very low-quality evidence studies prone to bias.

### Implications for Research

More research is needed to generate high-quality evidence in low and moderate endemic settings regarding the effects of house improvements in different endemic settings. Specifically, holistic house improvements to modern standards and not just components of house improvements like screening, closing eaves or iron roofing as these can have other unintended consequences such as poor ventilation and higher indoor temperatures.

### Implications for Practice

In all malaria-endemic areas, house improvements may be one of the additional tools for policymakers and program managers to consider implementing in malaria programs.

## Authors’ Contributions

MN and OA discussed the concept, MN drafted the protocol, and OA reviewed and provided inputs and guidance. Both authors reviewed and approved the protocol for submission. MN Mukumbuta Nawa, OA Olatunji Adetokunboh.

## Supporting information

Search Strategy

## Data Availability

All data produced in the present work are contained in the manuscript.

## Acknowledgements

The authors wish to acknowledge Prof Peter Nyasulu and Ms Vera Ngah of Stellenbosch University. We also wish to thank Mrs Catherine Mupeyo Mudala and Mrs Sylvia Tembo Banda, who, during screening, data abstraction, risk of bias and grading of the studies.

## Patents and public involvement

No patients or the public were involvement

