## Supplementary material for "The Effects of Improved Housing on Malaria Transmission in Different Endemic Zones: A Systematic Review and Meta-Analysis": Table 1

**Table 1: Characteristics of Included Studies**

| **Author** | **Year** | **Continent** | **Country** | **Population** | **Endemicity** | **Design** | **Mal. Measure** | **Effect** | **Estimate** | **Lower CI** | **Upper CI** | **P value** | **Study Pop.** |
| --- | --- | --- | --- | --- | --- | --- | --- | --- | --- | --- | --- | --- | --- |
| Dahesh et al. | 2009 | Africa | Egypt | All age | Low | Case-control | Parasitaemia | Odds ratio | 0.68 | 0.28 | 1.66 |  |  |
| Dlamini et al. | 2017 | Africa | Swaziland | All age | Moderate | Cross-Sec | Parasitaemia | Odds ratio | 0.47 | 0.28 | 0.79 |  | 11426 |
| Ippolito et al. | 2017 | Africa | Zambia | All age | Moderate | Cross-Sec | Parasitaemia | Odds ratio | 0.26 | 0.09 | 0.73 | 0.01 | 2788 |
| Jatta et al. | 2018 | Africa | Gambia | Mosq. Pop | Moderate | RCT | Vector den | Mean Ratio | 0.33 | 0.22 | 0.51 | 0.001 | 4762 |
| Manyangadze | 2022 | Africa | Zimbabwe | All age | Low | Cross-Sec | Parasitaemia | Odds ratio | 0.41 | 0.2 | 0.83 |  |  |
| Mohan et al. | 2021 | Asia | India | All age | Moderate | Cross-Sec | Parasitaemia | Odds ratio | 0.91 | 0.77 | 1 | 0.01 | 70671 |
| Morakinyo et al. | 2018 | Africa | Nigeria | 5 - 59 m | High | Cross-Sec | Parasitaemia | Odds ratio | 0.71 | 0.56 | 0.93 | 0.01 | 6991 |
| Mosha et al. | 2020 | Africa | Tanzania | 0.5 - 14 y | High | Cross-Sec | Parasitaemia | Odds ratio | 0.27 | 0.13 | 0.54 | 0.001 | 6918 |
| Mosha et al. | 2020 | Africa | Tanzania | 0.5 - 14 y | High | Cross-Sec | Vector den | Odds ratio | 0.83 | 0.67 | 1.02 | 0.072 | 6918 |
| Mundagowa et al. | 2020 | Africa | Zimbabwe | All age | Moderate | Case-control | Parasitaemia | Odds ratio | 0.23 | 0.11 | 0.51 | 0.01 | 150 |
| Nawa et al. | 2020 | Africa | Zambia | 6 - 59 m | High | Case-control | Parasitaemia | Odds ratio | 0.33 | 0.11 | 0.99 | 0.047 | 720 |
| Nawa et al. | 2019 | Africa | Zambia | 6 - 59 m | High | Cross-Sec | Parasitaemia | Odds ratio | 0.4 | 0.34 | 0.71 | 0.001 | 19320 |
| Okiring et al. | 2019 | Africa | Uganda | Preg wom. | High | Cohort | Incidence | IRR | 0.76 | 0.69 | 0.87 | 0.001 | 753 |
| Pinder et al. | 2021 | Africa | Gambia | 0.5 - 13 y | Moderate | RCT | Incidence | IRR | 1.68 | 1.11 | 2.255 | 0.014 | 785 |
| Pinder et al. | 2021 | Africa | Gambia | 0.5 - 13 y | Moderate | RCT | Vector den | IRR | 0.78 | 0.53 | 1.15 | 0.21 | 785 |
| Rek et al. | 2018 | Africa | Uganda | 0.5 - 10 y | High | Cohort | HBR | IRR | 0.27 | 0.17 | 0.42 | 0.001 | 384 |
| Rek et al. | 2018 | Africa | Uganda | 0.5 - 10 y | High | Cohort | HBR | IRR | 0.52 | 0.36 | 0.73 | 0.002 | 384 |
| Rugnao et al. | 2019 | Africa | Uganda | 2 - 10 y | High | Cross-Sec | Parasitaemia | Prev Ratio | 0.88 | 0.81 | 0.97 | 0.0008 | 8834 |
| Salunkhe et al. | 2019 | Asia | India | All age | Moderate | Cross-Sec | Parasitaemia | Odds ratio | 0.72 | 0.39 | 1.35 |  | 4954 |
| Sikalima et al. | 2021 | Africa | Zambia | All age | High | Cross-Sec | Parasitaemia | Odds ratio | 0.38 | 0.2 | 0.73 | 0.01 | 282 |
| Smith et al. | 2021 | Africa | Namibia | All age | Moderate | Case-control | Parasitaemia | Odds ratio | 0.5 | 0.36 | 0.69 |  | 1411 |
| Snyman et al. | 2015 | Africa | Uganda | 0 - 24 m | High | Cohort | Incidence | IRR | 0.54 | 0.39 | 0.78 | 0.001 | 515 |
| Tusting et al. | 2016 | Africa | Uganda | Mosq. Pop | Low | Cohort | HBR | IRR | 0.53 | 0.4 | 0.69 | 0.001 | 124746 |
| Tusting et al. | 2016 | Africa | Uganda | All age | Low | Cohort | Incidence | IRR | 0.93 | 0.68 | 1.28 | 0.67 | 2399 |
| Tusting et al. | 2016 | Africa | Uganda | All age | Low | Cohort | Parasitaemia | Odds ratio | 0.51 | 0.36 | 0.71 | 0.001 | 3367 |
| Wolff et al. | 2001 | Africa | Malawi | 0 - 5 y | High | Cross-Sec | Parasitaemia | Odds ratio | 0.73 | 0.36 | 1.41 |  | 318 |
| Zaidi et al. | 2015 | Asia | Pakistan | All age | Moderate | Cross-Sec | Parasitaemia | Odds ratio | 0.65 | 0.39 | 1.1 |  | 1623 |

Note: y – years; m – months; mosq. Pop – mosquito population; preg. Wom – pregnant women; cross-sect – cross-sectional study; vector den – vector density; IRR – Incidence rate ratio
