## Supplementary material for "The Effects of Improved Housing on Malaria Transmission in Different Endemic Zones: A Systematic Review and Meta-Analysis": Table 2

Table 2: Risk of Bias in Included Studies

| **Risk of Bias in Randomised Controlled Trials** | | | | | | | | | | | |
| --- | --- | --- | --- | --- | --- | --- | --- | --- | --- | --- | --- |
| Authors | Year | Design | Allocation Sequence Generation | Allocation Concealment | Blinding of participants and Personelle | Blinding of Outcome Assessment | Incomplete Outcome Data | Selective Reporting | Others | Overall Assessment | Comments |
| Jatta et al. | 2018 | RCT | Low risk | Low risk | High risk | Low risk | Low risk | Low risk | Low risk | Some concerns | Open label |
| Pinder et al. | 2021 | RCT | Low risk | Low risk | High risk | Low risk | Low risk | Low risk | Low risk | Some concerns | Open label |
| **Risk of Bias in Observational Studies** | | | | | | | | | | | |
| Author | Year | Design | D1: Confounding | D2:Exposure measurement | D3:Participant Selection | D4:Post exposure interventions | D5: Missing data | D6: Measurement of outcome | D7: Reporting bias | Overall Rating | Comments |
| Dahesh et al. | 2009 | CC | Y | PY | N | N | No Info | N | N | Serious risk | Used unadjusted odds ratio |
| Dlamini et al. | 2017 | XS | PY | PY | N | N | No Info | N | N | Moderate risk |  |
| Ippolito et al. | 2017 | XS | PY | PY | N | N | No Info | N | N | Moderate risk |  |
| Manyangaze et al. | 2022 | XS | PY | PY | N | N | No Info | PY | N | Serious risk | Malaria cases were assessed over two decades. Recall bias |
| Mohan et al. | 2021 | XS | PY | PY | N | N | No Info | Y | N | Serious risk | High chance of recall bias of malaria in past two year |
| Morakinyo et al. | 2018 | XS | PY | PY | N | N | No Info | N | N | Moderate risk |  |
| Mosha et al. | 2020 | XS | PY | PY | N | N | No info | N | N | Moderate risk |  |
| Mundagowa et al | 2020 | CC | PY | PY | N | N | N | N | N | Moderate risk |  |
| Nawa et al. | 2020 | CC | PY | PY | N | N | N | N | N | Moderate risk |  |
| Nawa et al. | 2019 | XS | PY | PY | N | N | N | N | N | Moderate risk |  |
| Okiring et al | 2019 | CH | N | N | N | N | N | N | N | Low risk |  |
| Rek et al | 2018 | CH | N | N | N | N | N | N | N | Low risk |  |
| Rugnao et al | 2019 | XS | PY | PY | N | N | No Info | N | N | Moderate risk |  |
| Salunkhe et al | 2019 | XS | PY | PY | N | N | No Info | N | N | Moderate risk |  |
| Smith et al. | 2021 | CC | PY | PY | N | N | No Info | PY | N | Moderate risk |  |
| Snyman et al | 2015 | CH | N | N | N | N | No Info | N | N | Low risk |  |
| Tusting et al. | 2016 | CH | N | N | N | N | No Info | N | N | Low risk |  |
| Wolff et al. | 2001 | XS | PY | PY | PY | N | No Info | N | N | Serious risk | A committee selected improved house beneficiaries |
| Zaidi et al. | 2015 | XS | Y | PY | N | N | No info | N | N | Serious risk | Used unadjusted odds ratio |
| CC = Case-Control, XS = Cross-Sectional, CH = Cohort, Y = Yes, PY = Probably Yes, N= No, No Info = No Information | | | | | | | | | | | |
