## Supplementary material for "The Effects of Improved Housing on Malaria Transmission in Different Endemic Zones: A Systematic Review and Meta-Analysis": Table 3

**Table 3: Summary of Pooled Measures of Association**

| **No.** | **Study Design** | **No. of Studies** | **Endemicity** | **Outcome** | **Association Type** | **Measure (95% CI)** | **Heterogeneity Overall** | **Heterogeneity Subgroups** |
| --- | --- | --- | --- | --- | --- | --- | --- | --- |
| 1 | Cross-Sectional | 8 | All | Parasitaemia | aOR | 0.47 (0.31 - 0.72) | 0.79 | 0.90 |
|  |  | 4 | High | Parasitaemia | aOR | 0.43 (0.29 - 0.63) |  | 0.39 |
|  |  | 1 | Moderate | Parasitaemia | aOR | 0.91 (0.77 - 1.07) |  | - |
|  |  | 2 | Low | Parasitaemia | aOR | 0.42 (0.26 - 0.66) |  | 0.01 |
| 2 | Cross-Sectional | 5 | All | Parasitaemia | uOR | 0.67 (0.55 - 0.81) | 0 | 0.03 |
|  |  | 2 | High | Parasitaemia | uOR | 0.71 (0.58 - 0.90) |  | 0 |
|  |  | 2 | Moderate | Parasitaemia | uOR | 0.67 (0.46 - 0.99) |  | 0 |
|  |  | 1 | Low | Parasitaemia | uOR | 0.41 (0.20 - 0.83) |  | - |
| 3 | Case-Control | 2 | All | Parasitaemia | aOR | 0.52 (0.38 - 0.70) | 0 | 0 |
|  |  | 1 | High | Parasitaemia | aOR | 0.33 (0.11 - 0.99) |  | - |
|  |  | 0 | Moderate | Parasitaemia | aOR | - |  | - |
|  |  | 1 | Low | Parasitaemia | aOR | 0.54 (0.39 - 0.74) |  | - |
| 4 | Case-Control | 2 | All | Parasitaemia | uOR | 0.33 (0.06 - 1.75) | 0.71 | - |
|  |  | 0 | High | Parasitaemia | uOR | - |  | - |
|  |  | 0 | Moderate | Parasitaemia | uOR | - |  | - |
|  |  | 2 | Low | Parasitaemia | uOR | 0.33 (0.06 - 1.75) |  | 0.71 |
| 5 | Cohort | 2 | All | Incidence | aRR | 0.68 (0.48 - 0.96) | 0.71 | - |
|  |  | 2 | High | Incidence | aRR | 0.68 (0.48 - 0.96) |  | 0.71 |
|  |  | 0 | Moderate | Incidence | aRR | - |  | - |
|  |  | 0 | Low | Incidence | aRR | - |  | - |
| 6 | Cohort | 2 | All | Incidence | uRR | 0.89 (0.70 - 1.14) | 0 | - |
|  |  | 2 | High | Incidence | uRR | 0.89 (0.70 - 1.14) |  | 0 |
|  |  | 0 | Moderate | Incidence | uRR | - |  | - |
|  |  | 0 | Low | Incidence | uRR | - |  | - |
| 7 | Cohort | 2 | All | Parasitaemia | uRR | 0.63 (0.41 - 0.97) | 0.69 | - |
|  |  | 2 | High | Parasitaemia | uRR | 0.63 (0.41 - 0.97) |  | 0.69 |
|  |  | 0 | Moderate | Parasitaemia | uRR | - |  | - |
|  |  | 0 | Low | Parasitaemia | uRR | - |  | - |
| 8 | Cohort | 2 | All | Human Biting Rate | uRR | 0.53 (0.43 - 0.65) | 0 | - |
|  |  | 2 | High | Human Biting Rate | uRR | 0.53 (0.43 - 0.65) |  | 0 |
|  |  | 0 | Moderate | Human Biting Rate | uRR | - |  | - |
|  |  | 0 | Low | Human Biting Rate | uRR | - |  | - |

aOR = Adjusted Odds Ratio, uOR = Unadjusted Odds Ratio, aRR = Adjusted Risk Ratio, uRR = Unadjusted Risk Ratio
