## Supplementary material for "The Effects of Improved Housing on Malaria Transmission in Different Endemic Zones: A Systematic Review and Meta-Analysis": Table 4

Table 4: Quality of Evidence using the GRADE Approach

| **Certainty assessment** | | | | | | | | **№ of patients** | | **Effect** | | **Certainty** |
| --- | --- | --- | --- | --- | --- | --- | --- | --- | --- | --- | --- | --- |
| **Outcomes** | **No of Studies** | **Study design** | **Risk of bias** | **Inconsistency** | **Indirectness** | **Imprecision** | **Other considerations** | **Modern housing** | **Traditional housing** | **Relative**  **(95% CI)** | **Absolute**  **(95% CI)** |  |
| Parasitaemia  (Adjusted ORs) | 7 | observational  studies (cross-sectional) | very serious ^a^ | not serious ^b^ | not serious ^c^ | not serious ^d^ | none | 7188/74147 (9.7%) | 5273/28463 (18.5%) | **OR 0.47** (0.31 to 0.72) | **89 fewer per 1,000** (from 119 fewer to 45 fewer) | ⨁⨁◯◯ Low |
| Parasitaemia  (Unadjusted ORs) | 5 | observational studies (cross-sectional) | very serious ^a^ | serious ^e^ | not serious ^c^ | serious ^f^ | none | 595/4316 (13.8%) | 789/3224  (24.5%) | **OR 0.67** (0.20 to 0.83) | **66 fewer per 1,000** (from 184 fewer to 33 fewer) | ⨁◯◯◯ Very low |
| Parasitaemia  (Adjusted ORs) | 2 | observational studies (case-control) | very serious ^a^ | not serious ^b^ | not serious ^c^ | not serious ^d^ | none | 979 cases, | 1282 controls | **OR 0.52** (0.38 to 0.70) | - | ⨁◯◯◯ Very low |
|  |  |  |  |  |  |  |  | - | 31.0% |  | **121 fewer per 1,000** (from 164 fewer to 71 fewer) |  |
| Parasitaemia  (Unadjusted ORs) | 2 | observational studies (case-control) | very serious ^a^ | serious ^e^ | not serious ^c^ | serious ^g^ | none | 51 cases, | 80 controls | **OR 0.33** (0.06 to 1.75) | **-** | ⨁◯◯◯ Very low |
|  |  |  |  |  |  |  |  | - | 42.0% |  | **227 fewer per 1,000** (from 378 fewer to 139 fewer) |  |
| Incidence Rate Ratios (Adjusted) | 2 | observational studies (cohort) | serious ^h^ | not serious ^b^ | not serious ^c^ | serious ^f^ | none | 73/174 (42.0%) | 463/957 (48.4%) | **RR 0.68** (0.48 to 0.96) | **155 fewer per 1,000** (from 252 fewer to 19 fewer) | ⨁◯◯◯ Very low |

CI: confidence interval; OR: odds ratio; RR: risk ratio

a. Downgraded for very serious risk of bias: All studies were observational with a significant risk of bias.
b. Most studies observed a protective effect of modern housing compared to traditional housing.
c. No serious indirectness: These studies were conducted in different settings and countries. The findings are generalisable in other places.
d. No serious imprecision: The overall effect was statistically significant and clinically important.
e. Few studies observed a protective effect of modern housing compared to traditional housing.
f. The overall effect was not statistically significant.
g. The overall effect was not statistically significant and with wide confidence intervals.
h. Downgraded for serious risk of bias: All studies were observational and non-randomised.
