## Supplementary figures and images for "The Effects of Improved Housing on Malaria Transmission in Different Endemic Zones: A Systematic Review and Meta-Analysis"

### Figure 1

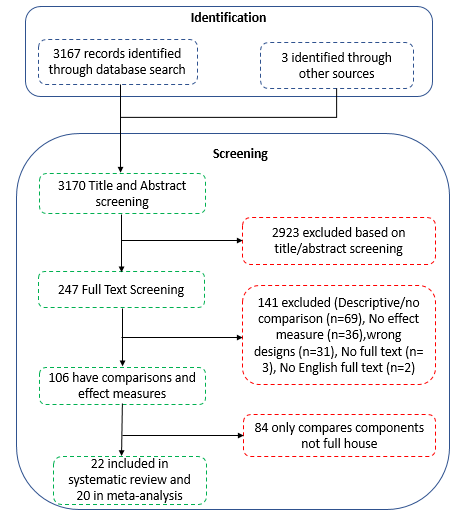

Figure 1: Inclusion Flow Chart
