## Supplementary material for "The Effects of Improved Housing on Malaria Transmission in Different Endemic Zones: A Systematic Review and Meta-Analysis": Search Strategy

**SEARCH STRINGS AND PROCESS**

**Search Output**

|  | **Database** | **No of publications** |
| --- | --- | --- |
| 1 | Scopus | 1860 |
| 2 | PubMed | 1918 |
| 3 | Medicus | 127 |
| 4 | Cochrane Library | 658 |
| 5 | Web of Science | 1233 |
|  | Total | 5796 |

After removal of duplicates with EndNote and Rayyan = 3171

Group 1 screening = 1586 (Nawa & Caroline)

Group 2 screening = 1585 (Nawa & Sylvia)

**SCOPUS**

1,860 document results

( TITLE-ABS-KEY ( malaria OR plasmodium OR anopheles OR "mosquito control" ) ) AND ( TITLE-ABS-KEY ( house OR houses OR housing OR home OR homes OR hut OR huts OR building OR buildings OR dwelling OR dwellings OR shelter OR shelters ) ) AND ( TITLE-ABS-KEY ( "randomised trial" OR "randomized trial" OR "randomised clinical trial" OR "randomized clinical trial" OR "randomised clinical trial" OR "randomized clinical trial" OR "cohort study" OR "longitudinal study" OR "cross-sectional survey" OR "cross sectional study" OR survey OR "case control study" OR "case-control study" ) ) AND ( LIMIT-TO ( PUBYEAR , 2022 ) OR LIMIT-TO ( PUBYEAR , 2021 ) OR LIMIT-TO ( PUBYEAR , 2020 ) OR LIMIT-TO ( PUBYEAR , 2019 ) OR LIMIT-TO ( PUBYEAR , 2018 ) OR LIMIT-TO ( PUBYEAR , 2017 ) OR LIMIT-TO ( PUBYEAR , 2016 ) OR LIMIT-TO ( PUBYEAR , 2015 ) OR LIMIT-TO ( PUBYEAR , 2014 ) OR LIMIT-TO ( PUBYEAR , 2013 ) OR LIMIT-TO ( PUBYEAR , 2012 ) OR LIMIT-TO ( PUBYEAR , 2011 ) OR LIMIT-TO ( PUBYEAR , 2010 ) OR LIMIT-TO ( PUBYEAR , 2009 ) OR LIMIT-TO ( PUBYEAR , 2008 ) OR LIMIT-TO ( PUBYEAR , 2007 ) OR LIMIT-TO ( PUBYEAR , 2006 ) OR LIMIT-TO ( PUBYEAR , 2005 ) OR LIMIT-TO ( PUBYEAR , 2004 ) OR LIMIT-TO ( PUBYEAR , 2003 ) OR LIMIT-TO ( PUBYEAR , 2002 ) OR LIMIT-TO ( PUBYEAR , 2001 ) OR LIMIT-TO ( PUBYEAR , 2000 ) OR LIMIT-TO ( PUBYEAR , 1999 ) OR LIMIT-TO ( PUBYEAR , 1998 ) OR LIMIT-TO ( PUBYEAR , 1997 ) OR LIMIT-TO ( PUBYEAR , 1996 ) OR LIMIT-TO ( PUBYEAR , 1995 ) OR LIMIT-TO ( PUBYEAR , 1994 ) OR LIMIT-TO ( PUBYEAR , 1993 ) OR LIMIT-TO ( PUBYEAR , 1992 ) OR LIMIT-TO ( PUBYEAR , 1991 ) OR LIMIT-TO ( PUBYEAR , 1990 ) OR LIMIT-TO ( PUBYEAR , 1989 ) OR LIMIT-TO ( PUBYEAR , 1988 ) OR LIMIT-TO ( PUBYEAR , 1987 ) )

**PUBMED**

| **Search** | **Query** | **Results** |
| --- | --- | --- |
| #5 | Search: #1 AND #2 AND #3 Filters: from 1987 - 2022 | [1,918](https://pubmed.ncbi.nlm.nih.gov/?term=%231+AND+%232+AND+%233&filter=years.1987-2022&sort=relevance) |
| #4 | Search: #1 AND #2 AND #3 | [1,953](https://pubmed.ncbi.nlm.nih.gov/?term=%231+AND+%232+AND+%233&sort=) |
| #3 | Search: "randomised trial" OR "randomized trial" OR "randomised clinical trial" OR "randomized clinical trial" OR "randomised clinical trial" OR "randomized clinical trial" OR "cohort study" OR "longitudinal study" OR "cross-sectional survey" OR "cross sectional study" OR survey OR "case control study" OR "case-control study" | [2,300,588](https://pubmed.ncbi.nlm.nih.gov/?term=%22randomised+trial%22+OR+%22randomized+trial%22+OR+%22randomised+clinical+trial%22+OR+%22randomized+clinical+trial%22+OR+%22randomised+clinical+trial%22+OR+%22randomized+clinical+trial%22+OR+%22cohort+study%22+OR+%22longitudinal+study%22+OR+%22cross-sectional+survey%22+OR+%22cross+sectional+study%22+OR+survey+OR+%22case+control+study%22+OR+%22case-control+study%22&sort=) |
| #2 | Search: house OR houses OR housing OR home OR homes OR hut OR huts OR building OR buildings OR dwelling OR dwellings OR shelter OR shelters | [942,980](https://pubmed.ncbi.nlm.nih.gov/?term=house+OR+houses+OR+housing+OR+home+OR+homes+OR+hut+OR+huts+OR+building+OR+buildings+OR+dwelling+OR+dwellings+OR+shelter+OR+shelters&sort=) |
| #1 | Search: malaria OR Plasmodium OR Anopheles OR "mosquito control" | [131,316](https://pubmed.ncbi.nlm.nih.gov/?term=malaria+OR+Plasmodium+OR+Anopheles+OR+%22mosquito+control%22&sort=) |

**COCHRANE LIBRARY**

658 Trials matching malaria OR Plasmodium OR Anopheles OR "mosquito control" in Title Abstract Keyword AND house OR houses OR housing OR home OR homes OR hut OR huts OR building OR buildings OR dwelling OR dwellings OR shelter OR shelters in Title Abstract Keyword - with Publication Year from 1987 to 2022, with Cochrane Library publication date Between Jan 1987 and Sep 2022, in Trials (Word variations have been searched)

**MEDICUS**

tw:((tw:(malaria OR plasmodium OR anopheles OR "mosquito control")) AND (tw:(house OR houses OR housing OR home OR homes OR hut OR huts OR building OR buildings OR dwelling OR dwellings OR shelter OR shelters)) AND (tw:("randomised trial" OR "randomized trial" OR "randomised clinical trial" OR "randomized clinical trial" OR "randomised clinical trial" OR "randomized clinical trial" OR "cohort study" OR "longitudinal study" OR "cross-sectional survey" OR "cross sectional study" OR survey OR "case control study" OR "case-control study"))) AND (year_cluster:[1987 TO 2022]) 127 documents

**WEB OF SCIENCES**

malaria OR Plasmodium OR Anopheles OR "mosquito control" (Topic) AND house OR houses OR housing OR home OR homes OR hut OR huts OR building OR buildings OR dwelling OR dwellings OR shelter OR shelters (Topic) AND "randomised trial" OR "randomized trial" OR "randomised clinical trial" OR "randomized clinical trial" OR "randomised clinical trial" OR "randomized clinical trial" OR "cohort study" OR "longitudinal study" OR "cross-sectional survey" OR "cross sectional study" OR survey OR "case control study" OR "case-control study" (Topic) 1233 documents
